# Circulating Micro-RNAs in Patients with Hypophosphatasia Results of the first micro-RNA analysis in HPP

**DOI:** 10.1101/2024.07.17.24310437

**Authors:** Judith Haschka, Zora Messner, Julia Feurstein, Benjamin Hadzimuratovic, Jochen Zwerina, Andreas B. Diendorfer, Marianne Pultar, Matthias Hackl, Martin Kuzma, Juraj Payer, Heinrich Resch, Roland Kocijan

## Abstract

**Introduction:** Adult hypophosphatasia (HPP) patients present with diffuse heterogenous symptoms often mimicking rheumatological diseases or osteoporosis and therefore accompanied by delayed diagnosis. The aim of this study was to identify circulating miRNAs in adult HPP patients and to identify potential associations with clinical patients’ characteristics.

**Methods:** We utilized untargeted miRNA biomarker discovery by small RNA-sequencing to investigate cell-free miRNA profiles in 24 adult HPP patients (pathogenic variant of the *ALPL* gene, HPP-related clinical symptoms and repeatedly low ALP) and 24 healthy controls.

**Results:** Patients and CTRL were comparable in age (47.9±14.2 vs. 45.9±8.8y, p=0.980) and sex (55.5% vs. 47.8% females, p=1.000). In total, 91% of patients reported musculoskeletal pain, 41% diffuse neurological symptoms and 64% history of fractures. In total, 84 miRNAs were significantly differently expressed between HPP and CTRLs in next generation sequencing (NGS) analysis(p<0.05). Of these, 14 miRNAs were selected (selection criteria: p<0.05, tissue specificity index >0.7, log_2_ FC >+0.8 or < −0.8) for validation using RT-qPCR, which verified 6 of 14 selected miRNAs (p<0.05; miR-122-3p, miR-140-5p, miR-143-3p, miR-155-5p, miR-451a, miR-92a-3p). Target prediction and enrichment analysis identified associations with the musculoskeletal system and the central nervous system. In total, 37 miRNAs correlated with ALP levels, but only three miRNAs with PLP (pyridoxal-5’- phosphate).

**Conclusions:** These findings highlight a profound involvement of multiple organ systems and the potential of miRNAs as biomarkers for the effect of HPP on various systems.

## Introduction

Hypophosphatasia (HPP) is a rare, inheritable disease caused by mutations of the *ALPL* gene, resulting in a deficiency of the tissue-non-specific isoenzyme of alkaline phosphatase (TNSALP).(1) To date more than 400 mutations have been identified, which can be transmitted autosomal dominantly or recessively.(2) TNSALP is present in numerous tissues, but in particular in mineralizing tissues like bone and teeth, the kidneys and the central nervous system (CNS).(3) Decreased activity of TNSALP leads to extracellular accumulation of its substrates inorganic pyrophosphate (PPi), pyridoxal 5’-phosphate (PLP) and phosphoethanolamine (PEA). Superabundance of extracellular PPi, a potent inhibitor of hydroxyapatite (HA) crystal formation and propagation, contributes to defective mineralization causing rickets and osteomalacia, respectively. Further, excessive PLP, the major circulating form of vitamin B6, reflects reduced PLP transport across the Blood Brain Barrier and cell membranes, resulting in altered metabolism and comprised neurotransmitter synthesis, including epileptic seizures especially in children.(4) Further, TNSALP plays an important role in purinergic signal transmission as ecto-nucleotidases to control ligand availability at purinergic nucleotide (P2) receptors and by generating extracellular nulceosides at adenosine (P1) receptors.(5) Severity and phenotype of HPP varies depending on the extent of changes in TNSALP and its substrates.(6, 7) Affected infants and children suffer from serious symptoms including deteriorations of the skeleton, joints, teeth and muscles, as well as involvement of the kidneys, brain and respiratory tract. Adults present with heterogenous diffuse symptoms like low-traumatic fractures, chronic muscle pain and weakness, arthropathy, dental abnormalities and recurrent headaches.(8–10) Therefore, HPP often mimics rheumatological diseases with the risk of misdiagnosis and delayed diagnosis.(11, 12) Diagnostic work-up is based on clinical findings, physical examination, radiographic or histopathological exams and laboratory abnormalities, including low activity of TNSALP and accumulation of its substrates as well as genetic testing.(4) However, diagnosis in adults can be challenging due to the unspecific symptoms and no diagnostic accuracy to identify patients at risk for fractures by standard methods like assessment of bone mineral density (BMD) by Dual X-Ray absorptiometry (DXA) or established bone turnover markers.(13, 14) The diversity of the clinical phenotypes in HPP highlights the need of additional diagnostic landmarks helping to identify patients with HPP. MicroRNAs (miRNAs) are short, single-stranded, non-coding RNAs which act as regulators of gene expression at post-transcriptional level and hereby modulate protein translation and synthesis.(15) They play a pivotal role in various biological processes, including bone homeostasis and differentiation of bone cells.(16) Several miRNAs have been identified as bone specific and their relevance in bone diseases has been demonstrated previously.(17) miRNAs have the potential to not only display metabolic processes but also to serve as potential biomarkers. No data on miRNAs in HPP patients are available to date. The aim of this study was to identify the circulating miRNA signature in adult HPP patients compared to healthy controls and to identify potential associations with clinical patients’ characteristics.

## Patients and Methods

### Patient population and Study Design

This cross sectional study enrolled clinically- and genetically-confirmed HPP patients and age and gender-matched healthy controls. HPP Patients meeting the following inclusion criteria were included into the study: i) pathogenic variant within the *ALPL* gene (class 4 or 5), ii) HPP-related clinical signs and symptoms and iii) repeated levels of alkaline phosphatase (ALP) below lower limit of normal. Other causes of low alkaline phosphatase levels were ruled out. Due to the rarity of the disease, no sample size calculation was carried out. The maximum number of patients who met the inclusion criteria were included. All patients were treatment-naïve for enzyme replacement therapy with asfostase alfa. Demographic and disease specific characteristics were obtained out of medical history or reported by patients (history of fractures, pseudofractures, impaired fracture healing, musculoskeletal pain, early tooth loss, chondrocalcinosis, nephrocalcinosis, neurological symptoms e.g. fatigue, headache or seizures, family history for HPP). Healthy controls had no chronic musculoskeletal pain, normal ALP levels and no past or present signs of musculoskeletal diseases. Healthy controls were sex- and aged matched. Serum samples for miRNA analyses of all patients and controls were obtained before 10 a.m. after over-night fasting, centrifugated and stored at −80°C. Routine laboratory results included C-reactive protein, blood count, calcium, phosphate, creatinine, gamma glutamyl transferase, alkaline phosphatase, ferritin, thyroid stimulating hormone and albumin. Additionally, PLP levels were assessed in serum. The study was conducted as a cooperation study of the Ludwig Boltzmann Institute for Osteology at Hanusch Hospital, the Comenius University Faculty of Medicine, University Hospital Bratislava and the St. Vincent Hospital Vienna. All patients and controls were investigated and serum samples collected at the two specialized Austrian centers for osteoporosis and bone diseases (St. Vincent Hospital and Hanusch Hospital) between 2017 and 2022. All patients provided written informed consent. The study was approved by the local ethical committees: Ethics Committee of the Vinzenz Group (201612_EK25) and Ethics Committee of the City of Vienna (EK-20-174-0820). Figure 1 illustrates the study flow chart.

**Figure 1:**
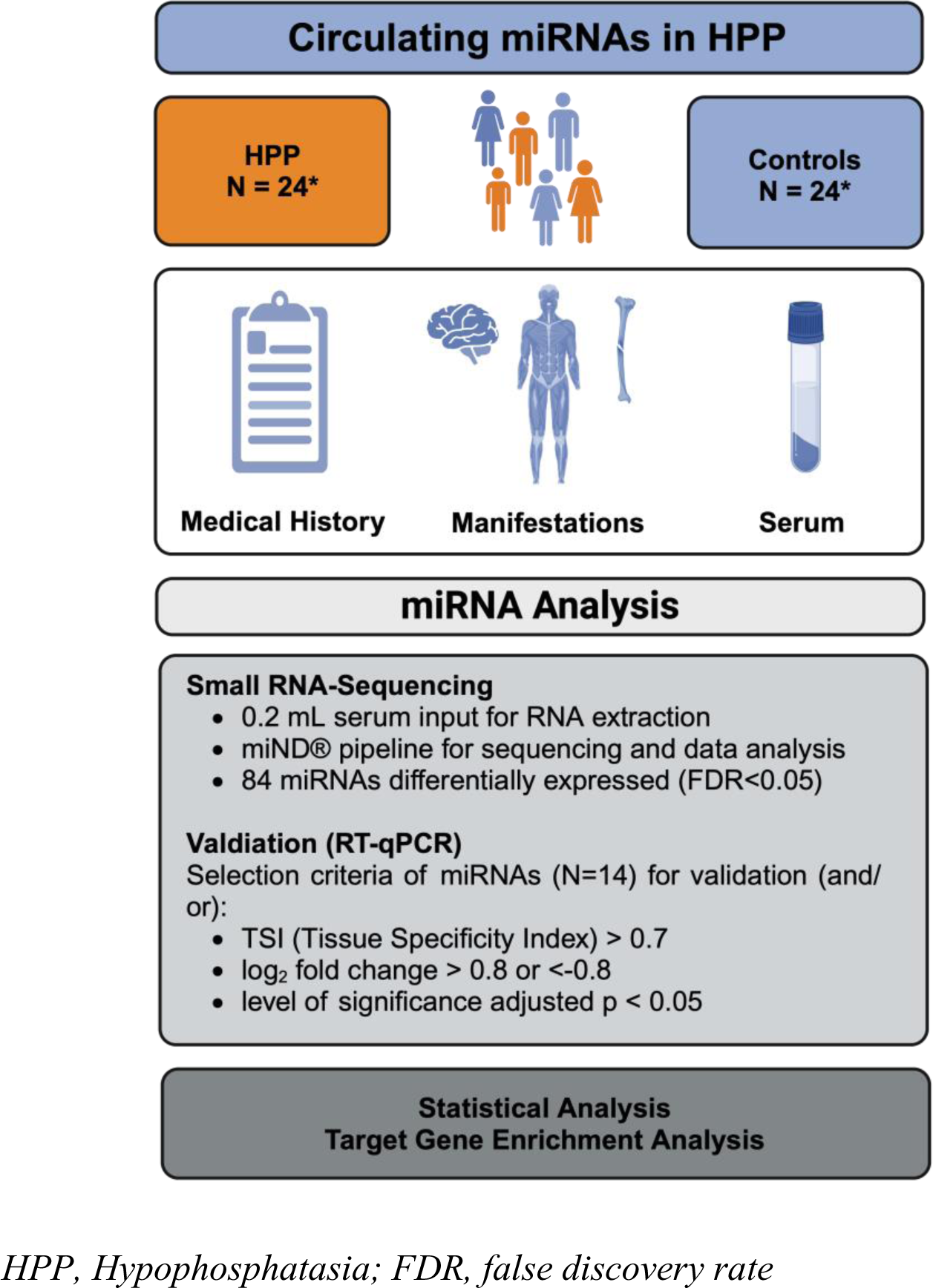
Study Flow chart of miRNA analysis in HPP patients and controls.

### miRNA analysis

#### Total RNA Extraction

total RNA was extracted from precisely 200 µl of serum using the Maxwell RSC miRNA tissue kit (AS1460, Promega). To monitor RNA extraction efficiency, miRCURY spike-in controls (Qiagen) were added to the lysis buffer prior to the extraction. To reduce the risk that excess amounts of miRCURY spike-ins interfere with sequencing depth for endogenous RNAs including miRNAs, pre-dilution of 1:250 was performed compared to the manufactureŕs instructions. Total RNA was eluted from beads in 50 µl nuclease-free water.

#### Small RNA-sequencing analysis

to construct small RNA-sequencing libraries a single-adapter ligation protocol (RealSeq Biosciences) was used as described previously.(18) Briefly, 8.5 µl of total RNA obtained from serum were mixed with 1 µl of miND® spike-in controls (TAmiRNA) and used as input for library construction following the manufacturer’s recommendations. Libraries were size purified to enrich for miRNA inserts on a 3% Agarose cassette using automated preparative gel electrophoresis (Bluepippin, Sage Biosystems), normalized to uniform molar concentration, pooled and sequencing on a NovaSeq 6000 SP1 flow-cell (Illumina). Raw data were processed using the miND® pipeline as described previously (19), to obtain microRNA read counts, which were further used as input for differential expression analysis with edgeR v3.32 (20) used the quasi-likelihood negative binomial generalized log-linear model functions provided by the package. The independent filtering method of DESeq2 (21) was adapted for use with edgeR to remove low abundant miRNAs and thus optimize the false discovery rate (FDR) correction to account for multiple testing.

#### Reverse-transcriptase quantitative PCR (RT-qPCR)

Selection of miRNAs for validation was based on the following criteria: i) tissue specificity index (TSI (22) > 0.7, ii) level of significance p < 0.05 and/or iii) logarithmic fold change (LOG FC) > +0.8 or < −0.8. Universal reverse transcription was performed using 2 µl total RNA from serum together with the miRCURY RT kit (Qiagen) in a 10 µl reaction volume at 42°C for 60 minutes followed by 5 minutes denaturation at 95°C. Cel-miR-39 was added to the RT mix to control for enzyme inhibition. cDNA was stored at −20°C until use for qPCR analysis. miRCURY LNA assays together with the miRCURY SYBR® Green mix (Qiagen) were used for amplification of target miRNAs in 96-well plate format on a Roche LC480 II. A total of 45 PCR cycles was performed using the cycling conditions recommended by the manufacturer. UniSp2 and cel-miR-39 (Qiagen) were detected to monitor RNA extraction and RT efficiency, respectively. Cq-values were obtained from Roche LC 480 software by applying the second derivative maximum method. Cq-values were further standardized to spike-in controls (UniSp2) to reduce the analytical variability. RT-qPCR data were further analyzed for group differences using non-parametric Kruskal-Wallis tests together with Benjamini-Hochberg adjustments for multiple testing.

### Statistical Analysis

Means of continuous clinical variables were compared between groups of categoric variables to analyze group homogeneity. Anderson-Darling’s test was used to assess normality. Depending on this result either an ANOVA (followed by t-tests) or a Kruskal-Wallis test (followed by Wilcoxon test) was used. Spearman correlation and rank-sum tests were used for correlation analysis of miRNA and other parameters. All p-values were adjusted for multiple testing according to the method by Benjamini-Hochberg.

Results are presented in mean±SD or median (IQR) depending on normality, if not stated otherwise.

### Target Gene Enrichment Analysis

Target genes for the subset of miRNAs were derived using the tool miRNAtap (23). Based on the combination of data obtained from different miRNA-mRNA interaction databases, the rank of predicted targets is aggregated from the 5 most commonly cited prediction algorithms: DIANA (24), Miranda (25), PicTar (26), TargetScan (27) and miRDB (28). The miRNA∼gene interaction has to be reported in at least 3 out of the 5 databases to be accounted for further enrichment analysis. The list of target genes was used to identify potentially enriched biological processes (BP). The tool topGO (29) with the org.Hs.eg.db (v3.18.0) was used. Fisher test was performed using all annotated human genes in the gene ontology dataset as background reference. The top 30 findings in total and after keyword specific filtering (“bone”/”osteo” and “neuro”) were obtained of significant results (p-value < 0.05).

## Results

### Patient characteristics

In total, 24 HPP patients and 24 controls (CTRL) were investigated. Two HPP patients and one control were excluded after serum samples did not pass quality control in miRNA analysis process. Table 1 summarizes demographic and clinical characteristics of the investigated study population. Patients and CTRL were comparable in age (47.9±14.2 vs. 45.9±8.8 years, p=0.980), sex (55.5% vs. 47.8% females, p=1.000) and BMI (25.4 (7.1) vs. 24.5 (2.7), p=0.980). ALP levels were lower in HPP patients (U/L; 23 (11) vs. 54 (28), p<0.0001) and 55% of patients had elevated PLP levels. Routine laboratory parameters were within normal range in both groups, with higher levels of calcium and phosphate in HPP patients. All HPP patients had a documented *ALPL* gene mutation, three of them had two heterozygous mutations and 36% of patients had a positive family history for HPP. In total, 64% of HPP patients reported a history of fractures, two of them had pseudofractures, one patient recurrent metatarsal fractures and one patient reported poor fracture healing. Chronic musculoskeletal pain was reported by 91% of patients and 77% had unspecific skeletal diseases in medical history e.g. tendinosis calcarean or calcification of the anterior spinal ligament. 41% of patients reported diffuse neurological symptoms e.g. headache or neuropathic pain. Demographic data of the study cohort are shown in table 1.

**Table 1.** Demographic data of the study cohort.

|  | **HPP**<br>**N=22<sup>†</sup>** | **CTRL**<br>**N=23<sup>†</sup>** | **P** |
| --- | --- | --- | --- |
| **Age [years]** | 47.9 ± 14.2 | 45.9 ± 8.8 | 0.980 |
| **Sex [females/males]** | 12/10 | 11/12 | 1.000 |
| **Height [cm]** | 168.1 ± 10.4 | 171.4 ± 9.9 | 0.726 |
| **Weight [kg]** | 75.0 ± 21.8 | 75.6 ± 12.0 | 0.980 |
| **BMI [kg/m<sup>2</sup>]** | 25.4 (7.1) | 24.5 (2.7) | 0.980 |
| **Laboratory Results** |  |  |  |
| **CRP [mg/L]** | 1.6 (6.4) | 1.0 (1.7) | 0.156 |
| **Hemoglobin [g/dL]** | 14.5 ± 1.4 | 14.4 ± 1.1 | 0.980 |
| **Leukocytes [G/L]** | 7.4 ± 2.1 | 6.5 ± 1.5 | 0.519 |
| **Calcium [mmol/L]** | 2.4 ± 0.1 | 2.3 ± 0.1 | **0.002** |
| **Phosphate [mmol/L]** | 1.2 ± 0.2 | 1.0 ± 0.2 | **0.0004** |
| **Creatinine [mg/dL]** | 0.8 (0.2) | 0.8 (0.1) | 0.747 |
| **GGT [U/L]** | 24 (16) | 23 (25) | 0.980 |
| **ALP [U/L]** | 23 (11) | 54 (28) | **< 0.0001** |
| **Ferritin [ng/mL]** | 140 (74) | 63 (57) | 0.160 |
| **TSH [mIU/L]** | 1.1 (0.9) | 1.0 (0.6) | 0.980 |
| **Albumin [g/L]** | 4.5 ± 0.3 | 4.5 ± 0.3 | 0.984 |
| **PLP [mcg/L]** | 36.3 (98.2) | - | - |
| **Min - Max** | 20.2 - 3000 | - | - |
| **PLP elevation, n (%)** | 12 (55) | - | - |
| **Family History & Clinical Symptoms** |  |  |  |
| **Family History of HPP, n (%)** | 8 (36) | - | - |
| **Muskuloskeletal Symptoms, n (%)** | **17 (77)** | - | - |
| History of Fractures, n (%) | 14 (64) | - | - |
| Pseudofractures, n (%) | 2 (9) | - | - |
| Recurrent Metatarsal Fractures, n (%) | 1 (5) | - | - |
| Poor Fracture Healing, n (%) | 1 (5) | - | - |
| Chronic Muskuloskeletal Pain, n (%) | 20 (91) | - | - |
| Muscular Cramps, n (%) | 11 (50) | - | - |
| Early Tooth Loss, n (%) | 7 (32) | - | - |
| **Development Disorder, n (%)** | **6 (27)** | - | - |
| **Nephrocalcinosis, n (%)** | **2 (9)** | - | - |
| **Diffuse neurological Symptoms, n (%)** | **9 (41)** | - | - |
| Seizures, n (%) | 0 (0) | - | - |
| **History of Pneumonia, n (%)** | **10 (46)** | - | - |

HPP, hypophosphatasia; CTRL, healthy controls; CRP, C-reactive protein; GGT, gamma-glutamyltransferase; ALP, alkaline phosphatase; TSH, thyroid-stimulating hormone; PLP, Pyridoxal 5'-phosphate in serum; <sup>†</sup>two patients and one control were excluded after quality control for miRNA analysis; all results after adjustment for multiple testing, level of significance p<0.05.

### miRNA analysis

Small RNA-sequencing data quality was assessed on the basis of i) sequencing depth, ii) miND® spike-in controls, iii) reads classification and iv) number of microRNAs detected (Supp Figure 1A-D). In total 3 samples were excluded from analysis, the remaining samples showed good spike-in performance, homogenous reads classification patterns, and on average >600 miRNAs above the limit of detection. After filtering of low abundant miRNAs, 281 were considered for statistical analysis, of which 84 miRNAs were significantly differently expressed between HPP and CTRLs in NGS analysis with 44 miRNAs being up-regulated, 40 miRNAs down-regulated. The volcano plot diagram in Figure 2 displays differences in miRNA expression levels. miRNA signature showed no sex-differences, two miRNAs showed significant correlation with age (miR-185-5p: r=-0.33, p=0.03; miR-206: r=0.44, p=0.003) and miR-93-3p was negatively correlated with BMI (r=-0.36 p=0.02).

**Figure 2:**
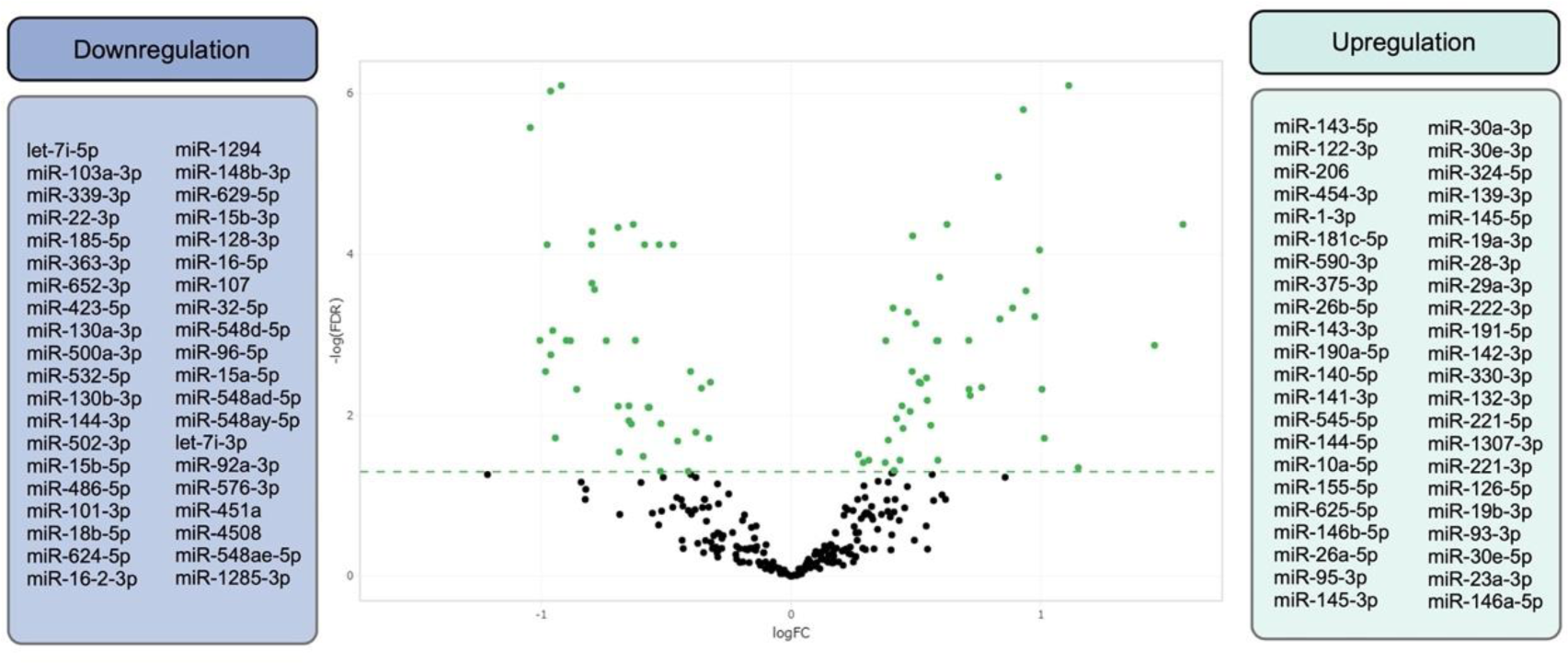
Differences of miRNA signatures between HPP and controls using Next-generation sequencing.

Based upon the predefined selection criteria for validation, 14 miRNAs (miR-140-5p, miR-451a, miR-92a-3p, miR-155-5p, miR-122-3p, miR-143-3p, miR-375-3p, miR-29a-3p, miR-146b-5p, miR-26b-5p, miR-144-3p, miR-1-3p, miR-107, miR-206) were further investigated. Validation analysis by RT-qPCR showed no signs of inhibition and homogenous recovery of both RNA and cDNA spike-in controls (Supp. Figure 2). Hence, RT-qPCR verified results for 6 out of 14 selected miRNAs being significantly different expressed between HPP patients and CTRL (adjusted p<0.05): miR-122-3p, miR-140-5p, miR-143-3p, miR-155-5p, miR-451a, miR-92a-3p (Figure 3). Further 5 out of 14 selected miRNAs showed a trend towards significance (adjusted p-value between 0.05 and 0.1): miR-375-3p, miR-29a-3p, miR-146b-5p, miR-26b-5p, miR-144-3p. All results of validation are presented in Table 2.

**Figure 3:**
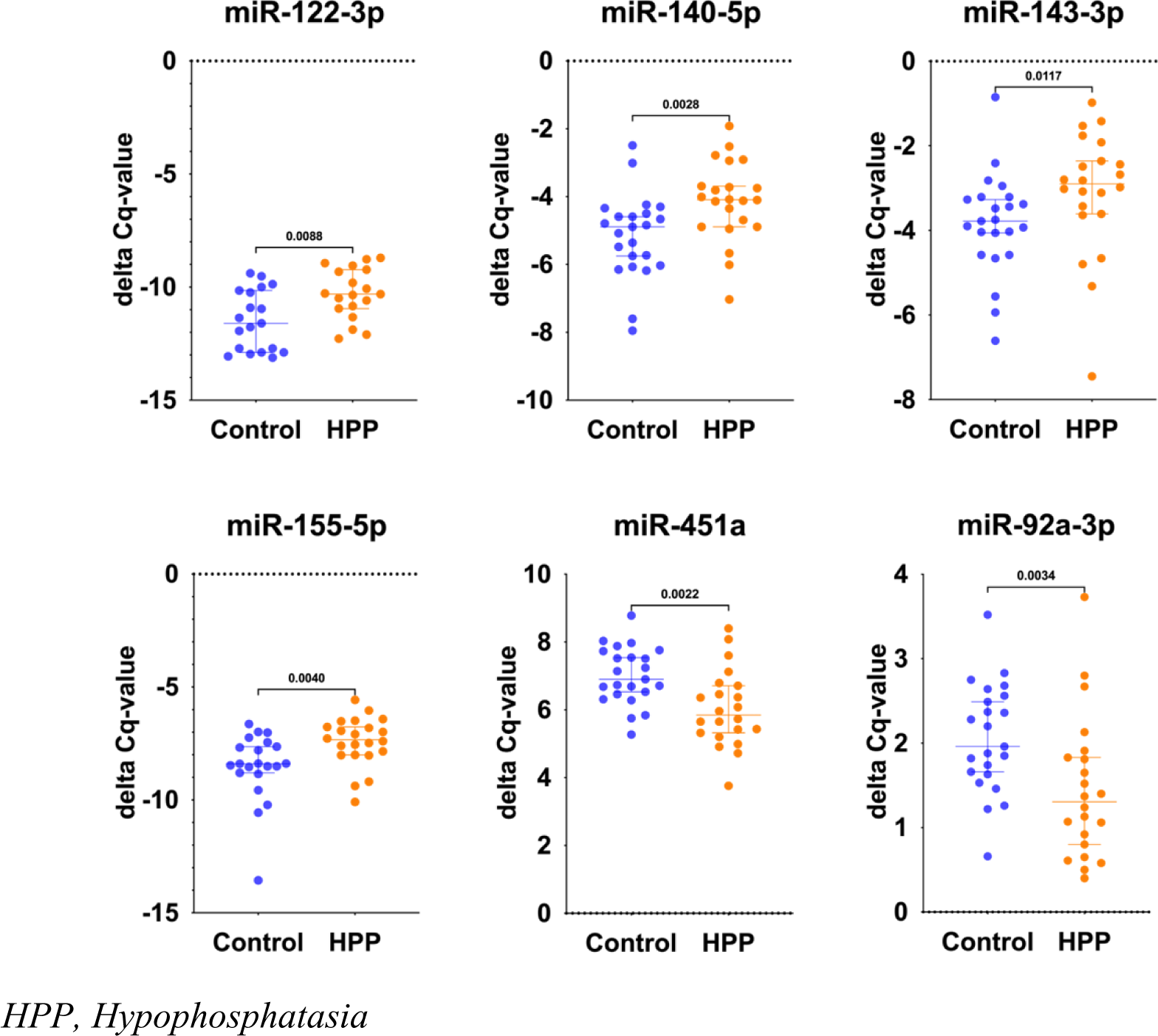
Differences of regulation levels of the top six miRNAs after validation using RT-qPCR.

**Table 2:**
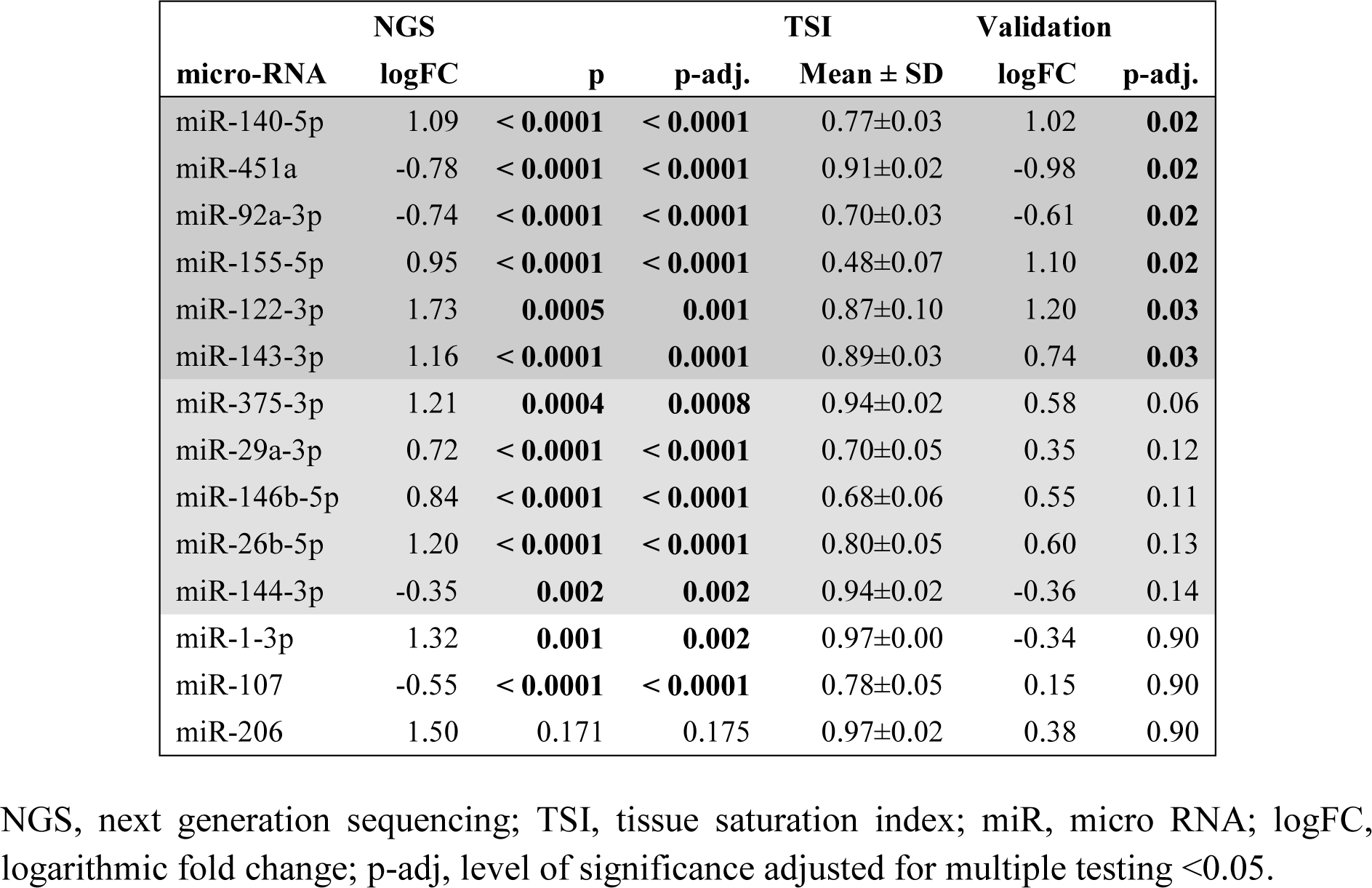
Results of miRNA analysis.

|  | NGS |  |  | TSI | Validation |  |
| --- | --- | --- | --- | --- | --- | --- |
| micro-RNA | logFC | p | p-adj. | Mean ± SD | logFC | p-adj. |
| miR-140-5p | 1.09 | < **0.0001** | < **0.0001** | 0.77±0.03 | 1.02 | **0.02** |
| miR-451a | -0.78 | < **0.0001** | < **0.0001** | 0.91±0.02 | -0.98 | **0.02** |
| miR-92a-3p | -0.74 | < **0.0001** | < **0.0001** | 0.70±0.03 | -0.61 | **0.02** |
| miR-155-5p | 0.95 | < **0.0001** | < **0.0001** | 0.48±0.07 | 1.10 | **0.02** |
| miR-122-3p | 1.73 | **0.0005** | **0.001** | 0.87±0.10 | 1.20 | **0.03** |
| miR-143-3p | 1.16 | < **0.0001** | **0.0001** | 0.89±0.03 | 0.74 | **0.03** |
| miR-375-3p | 1.21 | **0.0004** | **0.0008** | 0.94±0.02 | 0.58 | 0.06 |
| miR-29a-3p | 0.72 | < **0.0001** | < **0.0001** | 0.70±0.05 | 0.35 | 0.12 |
| miR-146b-5p | 0.84 | < **0.0001** | < **0.0001** | 0.68±0.06 | 0.55 | 0.11 |
| miR-26b-5p | 1.20 | < **0.0001** | < **0.0001** | 0.80±0.05 | 0.60 | 0.13 |
| miR-144-3p | -0.35 | **0.002** | **0.002** | 0.94±0.02 | -0.36 | 0.14 |
| miR-1-3p | 1.32 | **0.001** | **0.002** | 0.97±0.00 | -0.34 | 0.90 |
| miR-107 | -0.55 | < **0.0001** | < **0.0001** | 0.78±0.05 | 0.15 | 0.90 |
| miR-206 | 1.50 | 0.171 | 0.175 | 0.97±0.02 | 0.38 | 0.90 |

NGS, next generation sequencing; TSI, tissue saturation index; miR, micro RNA; logFC, logarithmic fold change; p-adj, level of significance adjusted for multiple testing <0.05.

### miRNA signature in HPP patients associated with serum ALP level and PLP elevation

In total, 37 miRNAs correlated significantly with ALP level, among those four miRNAs of the validated miRNAs using RT-qPCR (miR-140-5p r=-0.49, p=0.007, miR-451a r=0.52, p=0.002, miR-92a-3p r=0.39, p=0.05, miR-155-5p r=-0.53, p=0.002) and four miRNAs with a trend towards significance in validation (miR-29a-3p r=-0.41, p=0.05, miR-146b-5p r=-0.59, p=0.0001, miR-26b-5p r=-0.66, p <0.0001; miR-144-5p r=-0.59, p=0.0001). Three miRNAs correlated with PLP level (miR-29a-3p, miR-221-5p, miR-545-5p). Table 3 summarizes all significant correlations of miRNAs with ALP or PLP levels.

**Table 3:** Correlation of miRNA expression with ALP level and PLP level in HPP patients.

|  | miRNA | R | p-adj. | miRNA | R | p-adj. | miRNA | R | p-adj. |
| --- | --- | --- | --- | --- | --- | --- | --- | --- | --- |
| ALP<br>[U/L] | miR-16-5p | 0.65 | <**0.0001** | miR-32-5p | 0.52 | **0.002** | miR-30a-3p | -0.45 | **0.02** |
|  | miR-190a-5p | -0.65 | <**0.0001** | miR-451a | 0.52 | **0.002** | miR-142-3p | -0.44 | **0.03** |
|  | miR-26b-5p | -0.65 | <**0.0001** | miR-590-3p | -0.52 | **0.003** | miR-19a-3p | -0.45 | **0.03** |
|  | miR-144-5p | -0.59 | **0.0001** | miR-19b-3p | -0.51 | **0.003** | miR-10a-5p | -0.43 | **0.04** |
|  | miR-146b-5p | -0.59 | **0.0001** | miR-26a-5p | -0.51 | **0.004** | miR-145-3p | -0.43 | **0.04** |
|  | miR-625-5p | -0.60 | **0.0001** | miR-545-5p | -0.51 | **0.004** | miR-222-3p | -0.43 | **0.02** |
|  | miR-181c-5p | -0.57 | **0.0004** | miR-15a-5p | 0.50 | **0.004** | miR-221-3p | -0.42 | **0.02** |
|  | let-7i-3p | 0.57 | **0.0004** | miR-140-5p | -0.49 | **0.007** | miR-107 | 0.42 | **0.05** |
|  | miR-454-3p | -0.56 | **0.0006** | miR-128-3p | 0.48 | **0.01** | miR-30e-5p | -0.42 | **0.03** |
|  | miR-126-5p | -0.55 | **0.001** | miR-15b-3p | 0.48 | **0.01** | miR-29a-3p | -0.41 | **0.03** |
|  | miR-139-3p | -0.53 | **0.002** | miR-30e-3p | -0.48 | **0.01** | miR-92a-3p | 0.39 | **0.05** |
|  | miR-148b-3p | 0.53 | **0.002** | miR-191-5p | -0.46 | **0.02** |  |  |  |
|  | miR-155-5p | -0.53 | **0.002** | miR-145-5p | -0.46 | **0.02** |  |  |  |
| PLP<br>[mcg/L] | miR-29a-3p | -0.70 | **0.03** |  |  |  |  |  |  |
|  | miR-221-5p | -0.63 | **0.05** |  |  |  |  |  |  |
|  | miR-545-5p | -0.62 | **0.03** |  |  |  |  |  |  |

miRNA, micro-RNA; ALP, alkaline phosphatase level in serum; PLP, Pyridoxal 5'-phosphate level in serum; R, correlation coefficient; p-adj., level of significance adjusted for multiple testing p-adj. < 0.05.

### Association of miRNA signature and clinical characteristics in HPP patients

HPP patients with reported neurological symptoms showed a downregulation of miR-221-5p (p=0.02) and let-7i-3p (p=0.04), while the downregulation of miR-29a-3p and miR-122-3p lost level of significance after adjustment for multiple testing. In patients with reported musculoskeletal pain, miR-26b-5p (p=0.05) was significantly upregulated. Three more miRNAs were upregulated but lost level of significance after adjustment (miR-375-3p, miR-191-5p and miR-330-3p). In HPP patients with a history of fracture miR-4508 was significantly downregulated (p=0.05). In HPP patients with a positive family history of HPP two miRNAs remained significant (miR-339-3p and miR-130a-3p), while additional four miRNAs lost level of significance after adjustment, among those miR-92a-3p and miR-143-3p. In patients with reported early tooth loss, miR-23a-3p (p=0.04) and miR-590-3p (p=0.05) were aberrantly expressed.

### miRNA targets - Gene Ontology Enrichment Analysis

GO Analysis included more than 7000 GO terms. Overall, the most significant findings were directing to metabolic processes and RNA as well as DNA processing. Of all six significantly differently expressed miRNAs of the validation phase, an association with bone, the central nervous system and senescence, liver as well as inflammation has been previously described. GO terms specifically addressing the pathologies showed a total of 14 GO terms including “bone” or “osteo” (e.g. resorption, remodeling, mineralization, differentiation) and 51 GO terms included “neuro” or “myelin” or “membrane potential” (e.g. regulation of neurotransmitter receptor activity, neuronal action potential, central nervous system differentiation, regulation of membrane potential). Figure 4 shows blotting diagrams of the most significant findings as well as results of the specific GO terms. No significant associations were found with the GO terms “ATP”, “purinergic”, “senescence” or “aging”. (results not shown)

**Figure 4:**
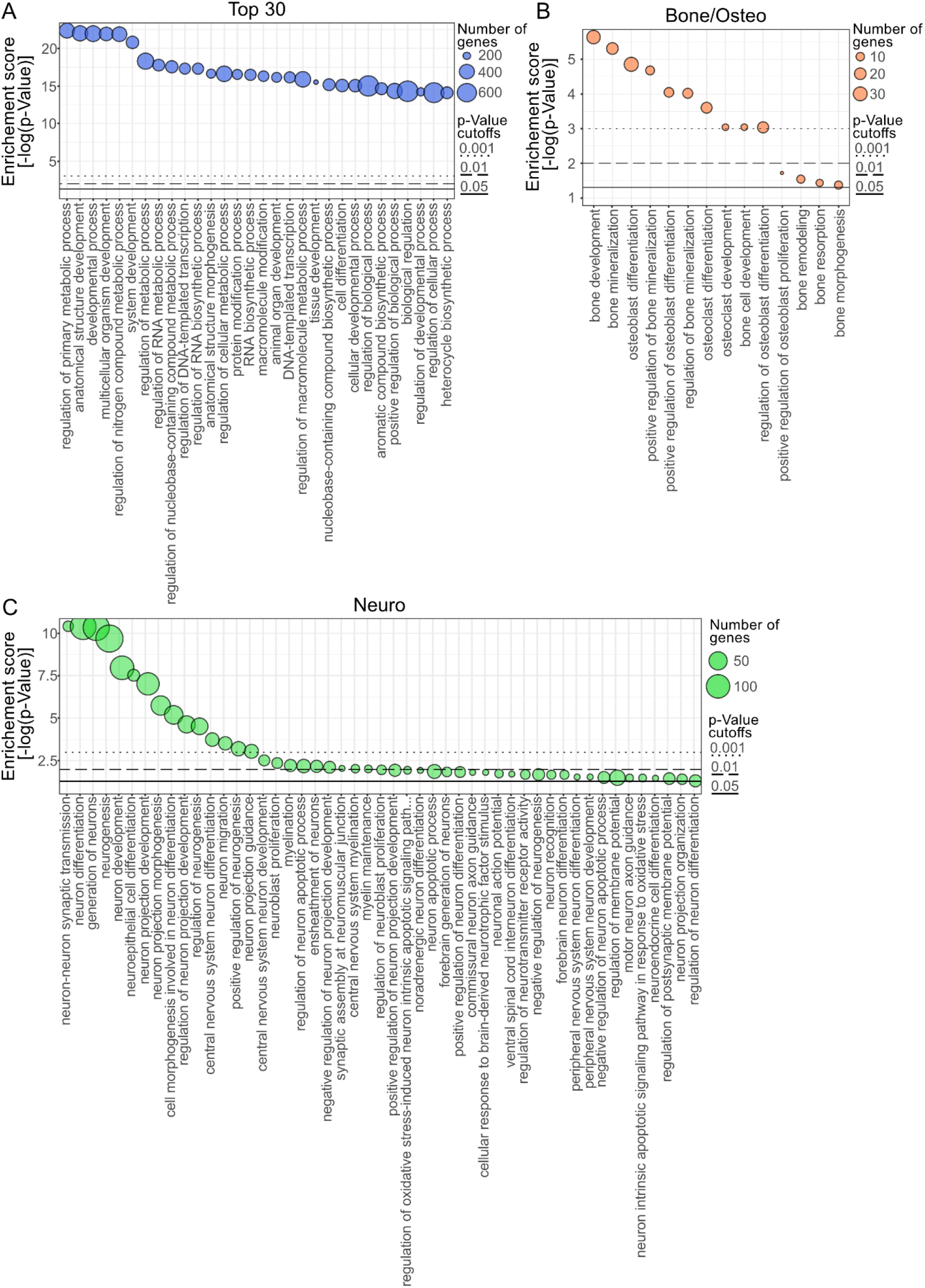
Target Gene Enrichment Analysis.

## Discussion

In this cross-sectional study, circulating miRNAs were analyzed in adult patients with genetically confirmed HPP. In total 84 miRNAs were differently expressed in HPP patients compared to controls, highlighting not only a profound but also widespread different miRNA profile of HPP patients. Adult HPP patients present with various skeletal and non-skeletal symptoms due to an inherited TNSALP deficiency. Therefore, the distinct aberrant miRNA signature of adult HPP underlines the involvement of multiple organ systems in this rare disease.

Established diagnostic procedures for bone assessment such as DXA, HR-pQCT or serum bone turnover markers are not specific, and thus not useful for HPP diagnosis, follow-up or fracture risk prediction.(8, 13, 14, 30) miRNAs have been the subject of intensive research in recent years and represent promising biomarkers for numerous diseases. Bone-specific miRNAs were already identified. In the present study, several of these miRNAs were significantly differently expressed in adult HPP patients and correlated with ALP levels and clinical symptoms. Six miRNAs could be validated by RT-qPCR (miR-143-3p, miR-122-3p, miR-140-5p, miR-155-5p, miR-451a and miR-92a-3p), making them top candidates for the discrimination between HPP and control. These miRNAs were reported to be associated not only to bone tissue, but also to cellular senescence and CNS.

MiR-143 acts as a direct suppressor on osteogenic differentiation via regulation of Osterix and was significantly upregulated in HPP patients.(31) Further, miR-122 has been previously reported to inhibit osteoblast proliferation and differentiation in ovariectomized rats and showed in vivo differences in expression level in osteoporotic patients.(32, 33) miR-451, one of the top regulated miRNAs in the present study, is a well-known regulator of osteogenesis and mineralization. Suppression of miR-451 was reported to inhibit bone loss in osteoporotic mice and was significantly lower in unhealed fractures and upregulated in newly formed tissue at fracture sites. Therefore miR-451 seems to play a pivotal role in fracture healing.(34, 35) This is an interesting point considering the impaired fracture healing in HPP patients.

The metabolic effects of TNSALP remain largely unexplored to date, but TNSALP plays a pivotal role in purinergic pathway by hydrolyzation of adenosine triphosphate (ATP), adenosine diphosphate (ADP) and monophosphate (AMP) to supply adenosine in various tissues.(5, 39) Differentiation and senescence of MSCs via ATP is orchestrated by alkaline phosphatase, playing therefore an important role in cellular function and ageing.(40) During cellular senescence in MSCs miR-140-5p is highly upregulated.(41) Further, a machine learning based study investigating cognitive aging identified three top ranked miRNAs for prediction of cognitive function (miR-140-5p, miR-197-3p, miR-501-3p), all of them linked to cellular senescence, inflammatory signals of atherosclerotic formation and potential development of neurodegenerative disorders e.g. alzheimers disease.(42) Therefore, the highly significant upregulation of miR-140-5p in HPP patients compared to controls is of special interest.

TNSALP also plays a pivotal role in the CNS. TNSALP is responsible for neuronal development and synaptic function by dephosphorylating PLP to membrane-permeable forms of vitamin B6 to allow diffusion across cell membranes. After re-phosphorylation it acts as an important coenzyme for biosynthesis for e.g. neurotransmitters, and regulator of purinergic transmission in the CNS.(39) In the Global HPP registry fatigue, generalized body pain and chronic muscle pain are frequent symptoms in adult HPP patients (23.4%, 22% and 19.1% respectively) regardless of age at HPP onset.(45) In a retrospective chart review of 82 adult and childhood-onset HPP patients, fatigue and diffuse neurological symptoms were reported in more than two thirds of patients besides musculoskeletal symptoms.(46) In the present cohort, 41% of patients reported diffuse neurological symptoms despite predominantly musculoskeletal symptoms. Several identified miRNAs including miR-92a-3p originate within the neurological system (48, 49) and correlated with ALP level. Additional, miR-29a-3p and miR-221 were differently expressed in patients with reported neurological symptoms and showed correlation with PLP level.

Further, miR-375-3p was upregulated in HPP, with a known negative regulatory effect on bone formation by inhibition of Wnt signaling pathway and showing response to osteoanabolic treatment with teriparatide in ovariectomized rats.(36, 37)

In the present study, the association to clinical symptoms was biased due to the cross-sectional design. However, miR-4508 which was significantly associated with the history of fractures, has been previously reported to show treatment response upon RANKL inhibition with Denosumab.(38) To conclude, in adult HPP patients several miRNAs with a known association to bone show a distinct pattern, qualifying them for further investigations.

On top of these miRNA results, gene ontology analysis gave further insight in the complexity of aberrant regulation in HPP patients. Despite the top findings of rather unspecific metabolic processes by the identified miRNAs, the more detailed search of affected organ systems highlighted a plethora of GO terms related with the central nervous system and bone.

This study has several strengths and limitations. miRNA analysis was performed in HPP patients for the first time. The aim of using NGS approach was to identify a broad variety of miRNAs of different organs and tissues. Due to the huge number of identified miRNAs in a rare patient population, a selection of 14 miRNAs, all of them previously reported with a known association with symptoms presented by adult HPP patients, were further validated using RT-qPCR. We verified 43% of selected miRNAs (p<0.05) and in total 79% with a p-value lower than 0.1, indicating a trend taking correction of multiple testing into account. Especially the three miRNAs without any trend of significance highlight the importance of validation using different analysis techniques. With respect to the small patient population in a rare disease cohort, validation in an independent cohort was not performed. Further, clinical characteristics were assessed out of medical history. No detailed structured assessment of neurological function or radiological assessment of the skeleton has been performed, may therefore be underreported and influence the correlation of symptoms and changes in miRNA profile. However, miRNAs have a promising potential and future analyses are warranted in different cohorts and during enzyme replacement treatment.

In conclusion, this cross-sectional study highlights significant differences of the miRNA signature in HPP patients and healthy controls. Among those, not only miRNAs with a known association to the musculoskeletal system, but also miRNAs associated with senescence, inflammation and the central nervous system have been identified. Adult HPP patients present with rather diffuse symptoms and the disease represents a multisystemic metabolic disease. Therefore, concise diagnostic standards are obligate and further investigation of potential new biomarkers warranted.

## Contributions

Conceptualization: Judith Haschka, Heinrich Resch, Roland Kocijan Data curation: Judith Haschka, Zora Messner, Julia Feurstein, Martin Kuzma, Juraj Payer, Jochen Zwerina, Heinrich Resch and Roland Kocijan; Methodology: Judith Haschka, Matthias Hackl, Roland Kocijan; Formal Analysis: Judith Haschka, Zora Messner, Benjamin Hadzimuratovic, Matthias Hackl, Andreas B. Diendorfer, Marianne Pultar and Roland Kocijan; Andreas B.Diendorfer, Marianne Pultar and Matthias Hackl performed miRNA analysis, validation, statistical analyses and Target Gene Enrichment Analysis; Funding Acquisition: Roland Kocijan, Heinrich Resch; Writing original draft: Judith Haschka, Zora Messner, Benjamin Hadzimuratovic and Roland Kocijan; Writing – review & editing: All authors critically revised and approved the final manuscript.

## Data Availability Statement

The data underlying this article will be shared on reasonable request to the corresponding author.

