## Supplemental Figure 1 and 2 for "Circulating Micro-RNAs in Patients with Hypophosphatasia Results of the first micro-RNA analysis in HPP"

Suppl. Figure 1: Data Quality Control of Next-Generation Sequencing

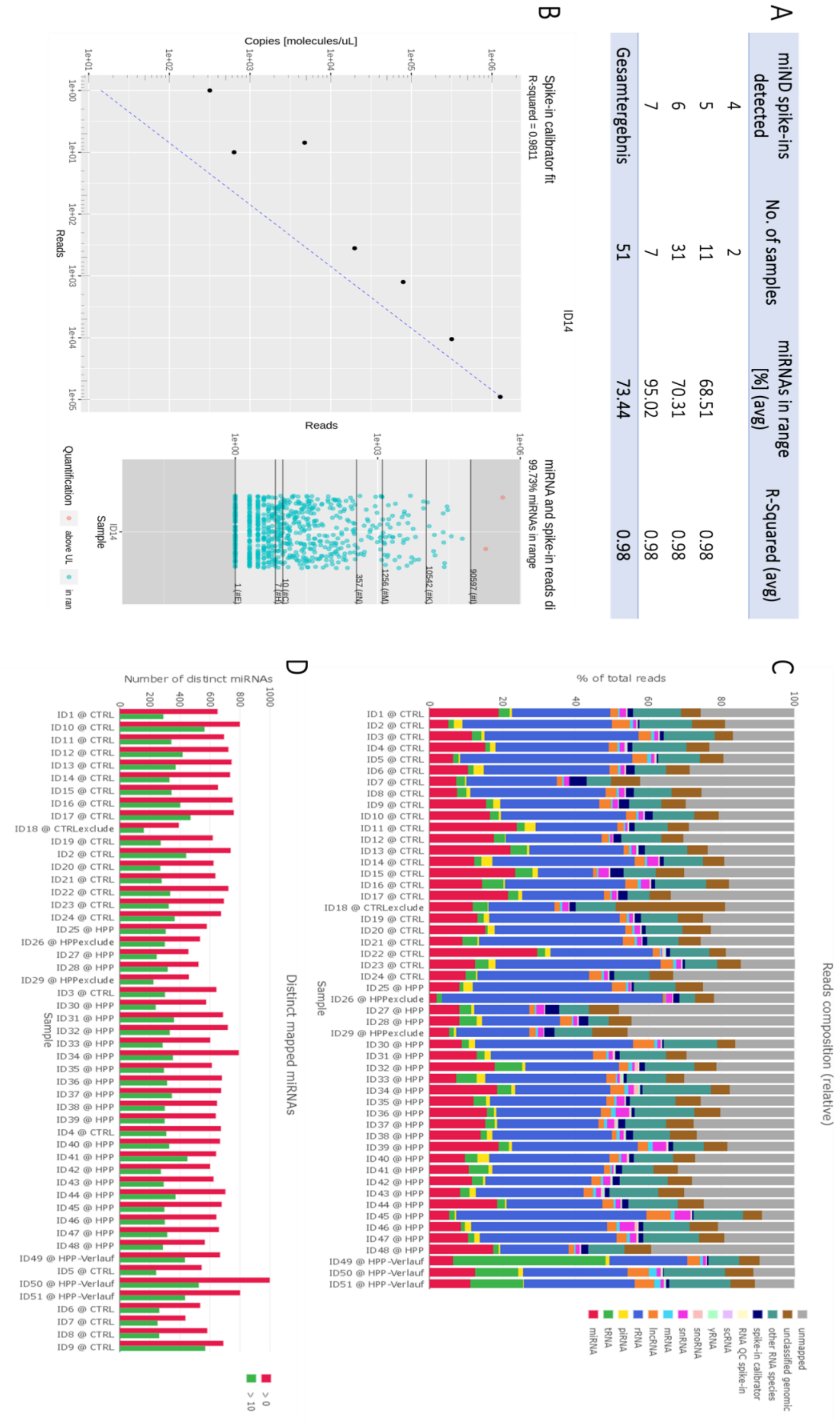


| miND spike-ins detected | No. of samples | miRNAs in range [%] (avg) | R-Squared (avg) |
|---|---|---|---|
| 4 | 2 | | |
| 5 | 11 | 68.51 | 0.98 |
| 6 | 31 | 70.31 | 0.98 |
| 7 | 7 | 95.02 | 0.98 |
| Gesamtergebnis | 51 | 73.44 | 0.98 |

Suppl. Figure 2: Data Quality Control of RT-qPCR

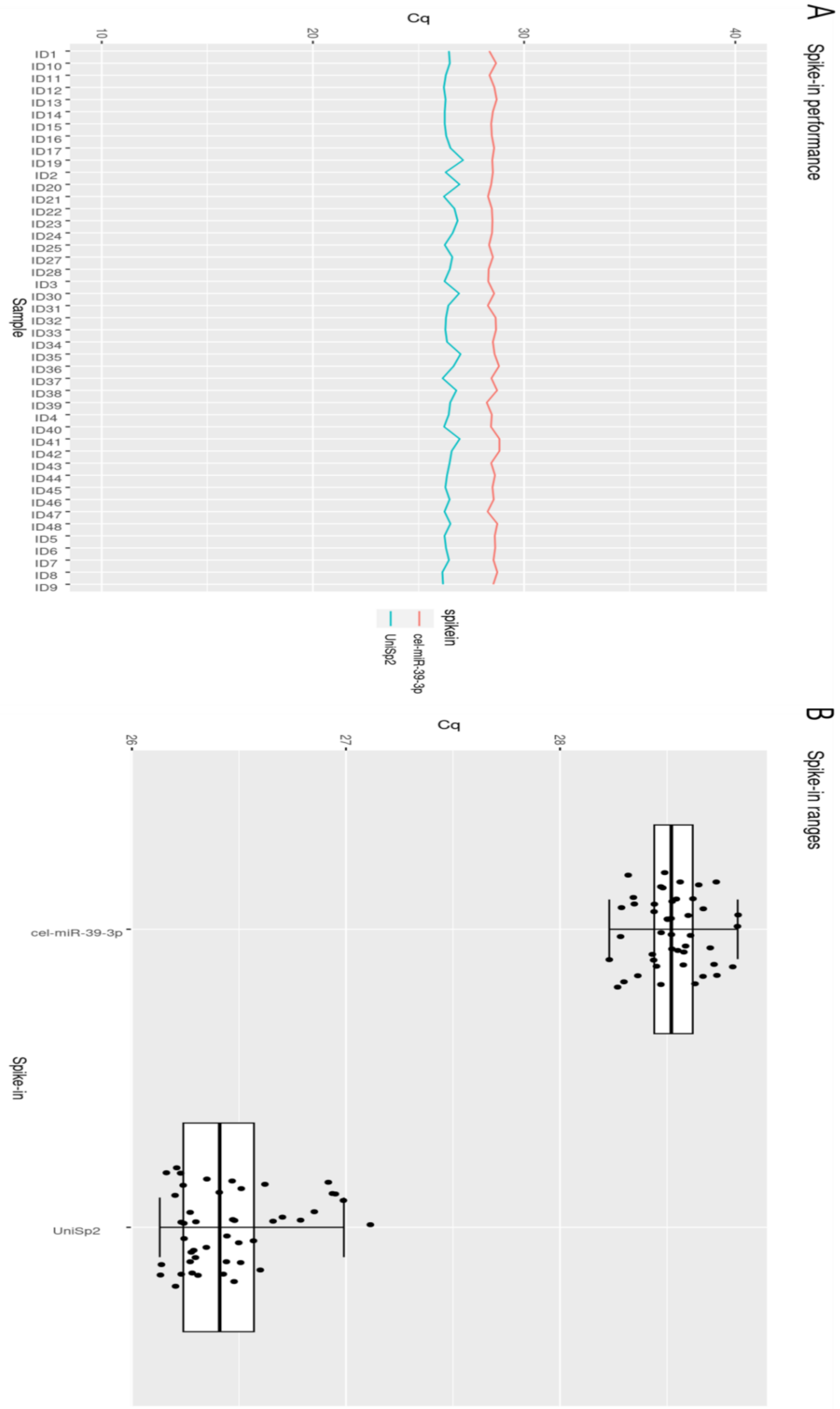
